# Lack of evidence for association of *UQCRC1* with Parkinson’s disease in Europeans

**DOI:** 10.1101/2020.09.04.20188243

**Authors:** Konstantin Senkevich, Sara Bandres-Ciga, Ziv Gan-Or, Lynne Krohn, on behalf of the International Parkinson’s Disease Genomics Consortium (IPDGC)

## Abstract

Recently, a novel variant p.Y314S in *UQCRC1* has been implicated as pathogenic in Parkinson’s disease (PD). In the current study, we aimed to examine the association of *UQCRC1* with PD in large cohorts of European origin. We examined common and rare genetic variation in *UQCRC1* using genome-wide association study data from the International Parkinson Disease Genomics Consortium (IPDGC), including 14,671 cases and 17,667 controls, and whole-genome sequencing data from the Accelerating Medicines Partnership - Parkinson’s disease initiative (AMP-PD), including 1,647 PD patients and 1,050 controls. No common variants were consistently associated with PD, and a variety of burden analyses did not reveal an association between rare variants in *UQCRC1* and PD. Therefore, our results do not support a major role for *UQCRC1* in PD in the European population, and additional studies in other populations are warranted.

## 1. Introduction

The genetics of Parkinson’s disease (PD) has been extensively studied over the last 20 years (Bandres-Ciga et al., 2020). A recent genome-wide association study (GWAS) identified 90 independent risk variants (Nalls et al., 2019); however, these variants explain less than 50% of the heritability of PD (Nalls et al., 2019), suggesting that other, unknown common and rare genetic variants affect the risk of PD. A novel variant p.Y314S in *UQCRC1* encoding the ubiquinol cytochrome c reductase core protein (UQCRC1) has been recently identified in five Taiwanese family members with parkinsonism by whole-exome sequencing (Lin et al., 2019). UQCRC1 is a mitochondrial protein and part of the respiratory chain III complex (Hoffman et al., 1993), which may play a role in mitochondrial respiration (Lin et al., 2019; Shan et al., 2019).

The purpose of this work is to examine the role of *UQCRC1* in a large-scale European PD population utilizing GWAS and whole-genome sequencing (WGS) data from the International Parkinson Disease Genomics Consortium (IPDGC) and Accelerating Medicines Partnership – Parkinson’s disease (AMP-PD) initiative.

## 2. Methods

The study populations included 14,671 PD patients and 17,667 controls from IPDGC, and 1,647 PD patients and 1,050 controls from AMP-PD (https://amp-pd.org/). Quality control of IPDGC GWAS data was performed on both individual and variant levels as previously described (Nalls et al., 2019). Similar quality control procedures were performed in the AMP-PD WGS data, as described by AMP-PD (https://amp-pd.org/whole-genome-data). We extracted *UQCRC1* genotyping data from both datasets using gene coordinates determined by UQCRC1 position (+/− 100kb): hg19: chr3:48,536,432-48,747,098; hg38: chr3:48,499,002-48,709,646. We utilized ANNOVAR (Wang et al., 2010) to annotate both data sets. PLINK 1.9 (Chang et al., 2015) was used for logistic regression (adjusted for age, sex and first 10 principal components) to study the association between common *UQCRC1* variants (with minor allele frequency [MAF] > 0.01) and PD in the IPDGC cohort. In the AMP-PD cohort, to study the burden of rare variants (defined as variants with MAF < 0.03 in the current data), a variety of methods were applied, including sequence Kernel association test and its optimized version (SKAT and SKAT-O), combined multivariate and collapsing (CMC), Zeggini and Madsen-Browning tests, as a part of the Rvtest package (Zhan et al., 2016). Bonferroni correction was applied to correct for multiple comparisons. All code used in the current study is available at our GitHub at https://github.com/ipdgc/IPDGC-Trainees/blob/master/UQCRC1_IPDGC_trainee.md.

## 3. Results

Using the IPDGC GWAS data, we identified 140 common variants in the selected region (Figure 1). None of the common variants annotated to *UQCRC1* were associated with PD (Supplementary Table 1). In the AMP-PD WGS data we identified 94 variants with MAF < 0.03 within or close to *UQCRC1* (Supplementary Table 2), including 9 nonsynonymous variants (Supplementary Table 2). We performed burden tests for three categories of variants: 1) all rare variants, 2) all rare coding variants and 3) all rare nonsynonymous variants. None of the burden tests showed association between rare *UQCRC1* variants and PD (Table 1). In both cohorts, we did not find the *UQCRC1* p.Y314S variant described in the original paper (Lin et al., 2019).

**Figure 1.**
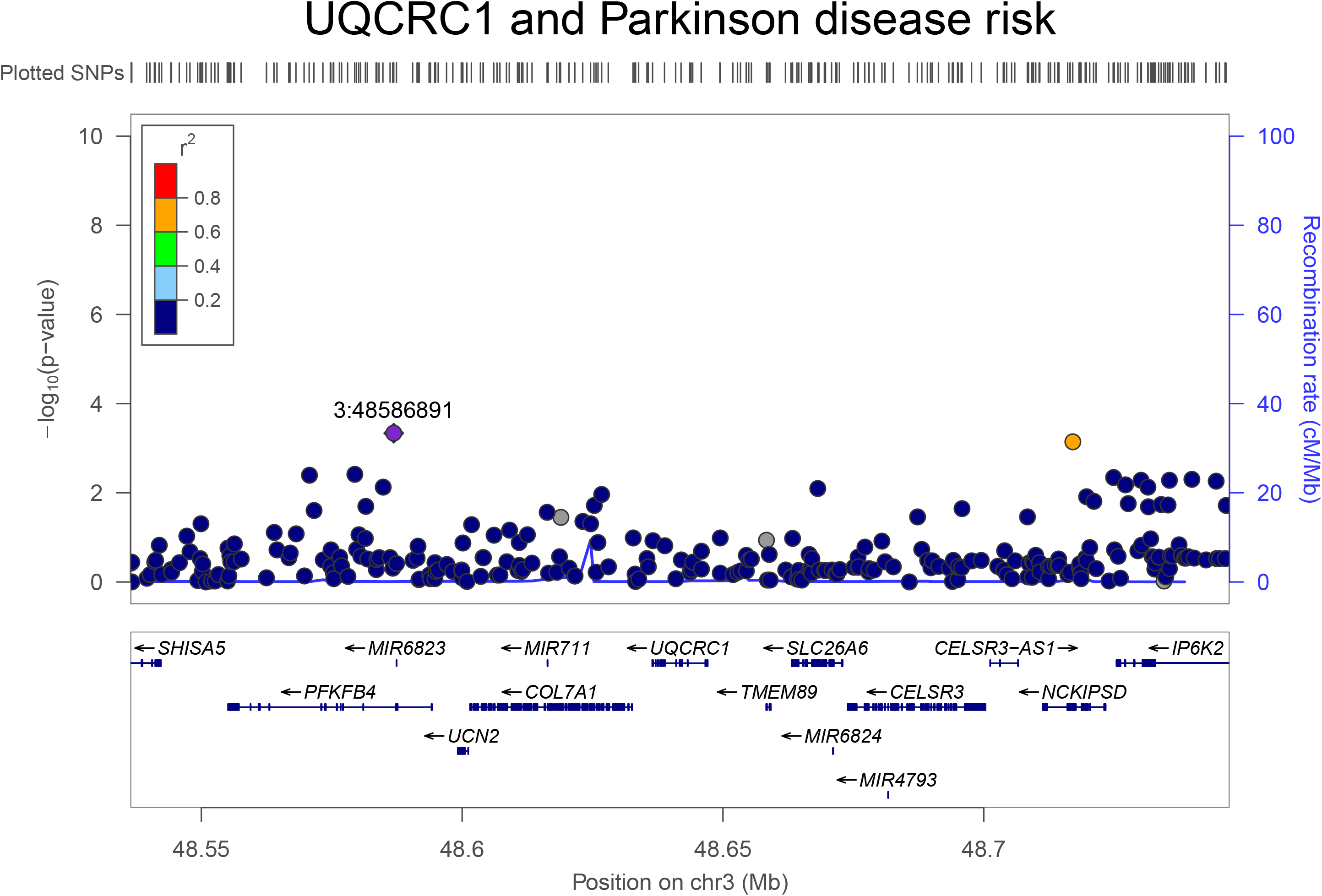
LocusZoom plot of IPDGC GWAS summary statistics showing variants with MAF more than 1% near the *UQCRC1* gene.

**Table 1.** Burden tests for *UQCRC1* in the AMP-PD cohort.

| Burden test | All variants |  |  | Coding variants |  |  | Nonsynonymous variants |  |  |
| --- | --- | --- | --- | --- | --- | --- | --- | --- | --- |
|  | Num Var | NumPolyVar | P-value | Num Var | NumPolyVar | P-value | NumVar | NumPolyVar | P-value |
| Zeggini | 73 | 54 | 0.562 | 18 | 13 | 0.619 | 12 | 9 | 0.721 |
| SKAT-O | 73 | 54 | 0.77 | 18 | 13 | 0.586 | 12 | 9 | 0.712 |
| SKAT | 73 | 54 | 0.865 | 18 | 13 | 0.476 | 12 | 9 | 0.594 |
| Madson-Browning | 73 | 54 | 0.487 | 18 | 13 | 0.046 | 12 | 9 | 0.109 |
| Fp | 73 | 54 | 0.712 | 18 | 13 | 0.489 | 12 | 9 | 0.898 |
| CMC | 73 | 54 | 0.596 | 18 | 13 | 0.629 | 12 | 9 | 0.731 |

SKAT- sequence Kernel association test; SKAT-O - Sequence Kernel association test optimized version; CMC - combined multivariate and collapsing; NumVar - number of variants; NumPolyVar - number of polymorphic genotypes;

## 4. Discussion

In the current study, we performed a comprehensive analysis of common and rare variants in *UQCRC1* using GWAS and WGS data from large cohorts of PD patients and controls.

In the original study in which *UQCRC1* variants were implicated, the authors described a family in which 5 carriers of *UQCRC1* p.Y314S had late-onset parkinsonism and axonal type sensorimotor polyneuropathy (Lin et al., 2019). Recent replication study in an eastern Chinese population did not reveal association between *UQCRC1* and PD (Lin et al., 2020).

It is possible that this variant is associated with a specific form of atypical parkinsonism, but not with typical PD. It has been shown that a number of genes previously reported as PD-associated (such as *DNAJC13, UCHL1, HTRA2, GIGYF2*, and *EIF4G1*) do not play a role in PD and thus, should not be regarded as PD genes (Foo et al., 2014; Krüger et al., 2011; Lesage et al., 2010; Saini et al., 2020). Furthermore, other genes (e.g. *ATP13A2, FBXO7*) that are associated with atypical forms of parkinsonism are often cited as PD-associated genes (Dehay et al., 2012; Deng et al., 2015; Weissbach et al., 2019). It is important to properly define which genes are involved in typical PD and which genes are not, particularly in the era of targeted drug development.

Overall, we did not find any evidence to support an important role for *UQCRC1* in PD patients of European origin. However, additional studies in other populations are required to further study the potential role of *UQCRC1* in PD, and we cannot rule out the possibility that very rare, specific *UQCRC1* variants are associated with PD or with atypical forms of parkinsonism.

## 5. Acknowledgements

We thank the participants for contributing to the study. We would like to also thank all members of the International Parkinson Disease Genomics Consortium (IPDGC). For a complete overview of members, acknowledgements and funding, please see http://pdgenetics.org/partners. This work was financially supported by grants from the Michael J. Fox Foundation, the Canadian Consortium on Neurodegeneration in Aging (CCNA), the Canada First Research Excellence Fund (CFREF), awarded to McGill University for the Healthy Brains for Healthy Lives initiative (HBHL), and Parkinson Canada. This research was supported in part by the Intramural Research Program of the NIH, National institute on Aging. KS is supported by a post-doctoral fellowship from the Canada First Research Excellence Fund (CFREF), awarded to McGill University for the Healthy Brains for Healthy Lives initiative (HBHL). ZGO is supported by the Fonds de recherche du Québec - Santé (FRQS) Chercheurs-boursiers award, in collaboration with Parkinson Quebec, and by the Young Investigator Award by Parkinson Canada.

## Data Availability

All code used in the current study is available at our GitHub.

https://github.com/ipdgc/IPDGC-Trainees/blob/master/UQCRC1_IPDGC_trainee.md

---

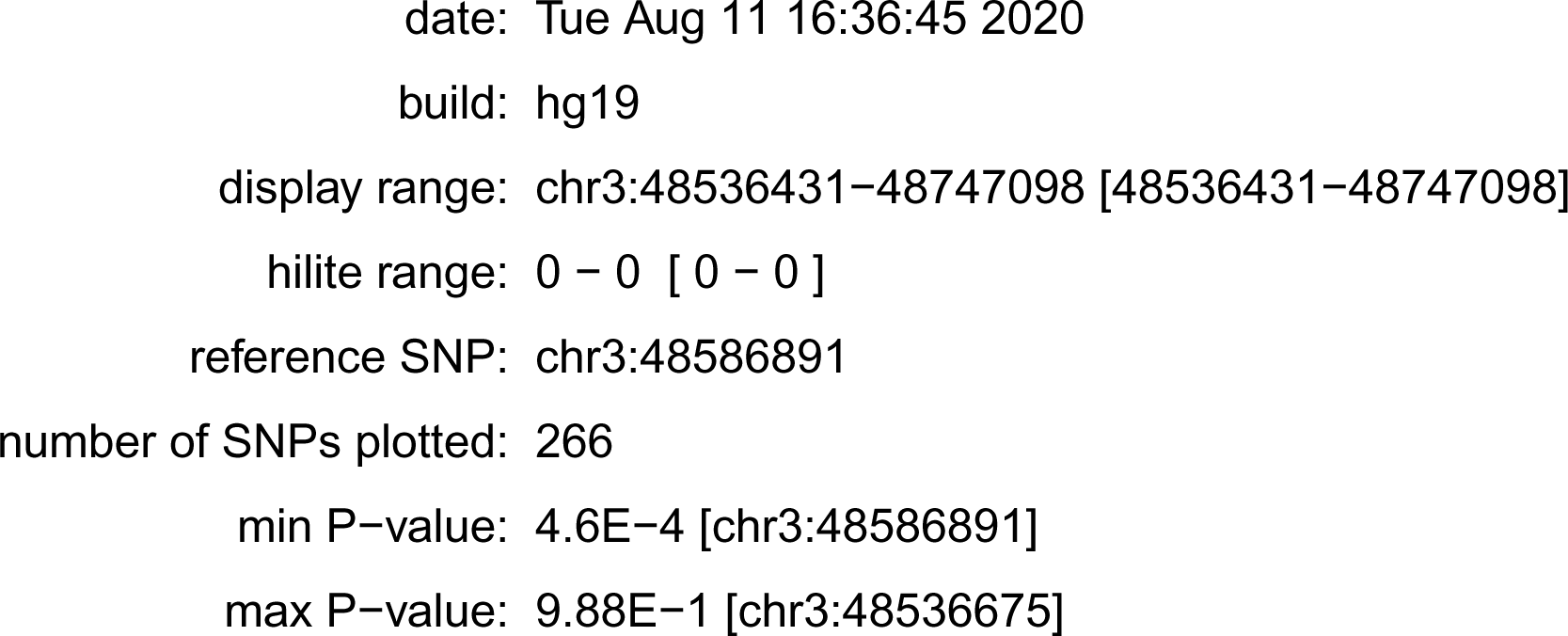

